# Use of pharmacy services in community-dwelling middle-aged and older adults; findings from The Irish Longitudinal Study on Ageing (TILDA)

**DOI:** 10.1101/2023.03.16.23287349

**Authors:** Logan T. Murry, Michelle Flood, Alice Holton, Rose Anne Kenny, Frank Moriarty

## Abstract

**Introduction:** The role of community pharmacists has evolved in recent years with expansion in pharmacy services offered. This study aims to assess pharmacy services use among adults aged ≥50 years in Ireland, and determine the demographic and clinical factors associated with pharmacy services use.

**Methods:** This cross-sectional study included community-dwelling participants in wave 4 of The Irish Longitudinal Study on Ageing (TILDA), aged ≥56 years who were self-respondents. TILDA is a nationally representative cohort study, with wave 4 data collected during 2016. TILDA collects participant demographics and health data, in addition to information on the use of several services when visiting the pharmacy in the last 12 months. Characteristics and pharmacy services use were summarised. Multivariate logistic regression was used to examine the association of demographic and health factors with reporting (i) any pharmacy service use and (ii) requesting medicines advice.

**Results:** Among 5,782 participants (55.5% female, mean age 68 years), 96.6% (5,587) reported visiting a pharmacy in the previous 12 months, and almost one fifth of these (1,094) availed of at least one specified pharmacy service. The most common non-dispensing services reported were requesting advice about medications (786, 13.6%), blood pressure monitoring (184, 3.2%), and vaccination (166, 2.9%). Controlling for other factors, female sex (odds ratio (OR) 1.32, 95%CI 1.14-1.52), third-level education (OR 1.85, 95%CI 1.51-2.27), higher rates of GP visits, private health insurance (OR 1.29, 95%CI 1.07-1.56), higher number of medications, loneliness, and respiratory condition diagnosis (OR 1.42, 95%CI 1.14-1.74) were associated with higher likelihood of availing of pharmacy services. The relationship between these factors and requesting medicines advice were similar.

**Conclusion:** A high proportion of middle-aged and older adults visit community pharmacy and a fifth avail of specified pharmacy services. Despite advances in the services offered in pharmacies, medicines advice remains at the core of pharmacists’ practice.

## Introduction

Community pharmacy has a potentially significant role in expanding access to community-based healthcare while providing enhanced services outside of traditional medication dispensing. In recent years, the role of community pharmacists has evolved globally, with expansion in a wide variety of pharmacy services offered in the community setting,(1) with further developments since the onset of the COVID-19 pandemic.(2-4)

In Ireland, enhanced or extended community pharmacy services include: interventions focused on smoking cessation, blood pressure monitoring, and inhaled medication adherence.(5) A recent study by Heinrich and Donovan identified that community pharmacists in Ireland reported confidence and willingness to play a greater role in medication deprescribing, but were limited in their involvement by time constraints while remuneration, interdisciplinary education, and bidirectional communication with prescribers facilitated community pharmacy deprescribing involvement.(6) When considering the extent to which pharmaceutical care is provided by community pharmacists, Ireland had a significantly higher total score than other European countries in the Behavioral Pharmaceutical Care Scale (BPCS), with the highest score for direct patient care activities.(7) These include patient assessment, implementation of therapeutic objectives and monitoring plans, and verifying patient understanding. Furthermore, there was a statistically significant positive relationship between provision of a health service (health screening; patient monitoring; domiciliary visiting; health promotion/education) and higher BPCS scores, suggesting community pharmacists and pharmacies in Ireland may be providing health services in greater number or frequency than other European countries.(7)

Although community pharmacies in Ireland have expanded enhanced and non-traditional service offerings, little is known about the uptake of pharmacy services in the middle-aged and older population in Ireland. As such, the objectives of this study were to 1) assess pharmacy service use among adults aged 50 years and over in Ireland and 2) determine what demographic and clinical factors are associated with pharmacy service use.

## Methods

This was a cross-sectional study of community-dwelling participants in wave 4 of The Irish Longitudinal Study on Ageing (TILDA). A nationally representative cohort study focusing on health, social and economic circumstances, TILDA recruited a sample of the population from a geo-directory of households in Ireland with residents 50 years of age and older with a baseline sample size of 8,175 participants and a 62% response rate.(8) Individuals younger than 50 years of age and residing in a nursing home or receiving institutional care were excluded from sampling. Wave 4 data was collected during 2016, and the current analysis included self-respondents (i.e. proxy respondents were omitted), who were all aged ≥56 years. Ethical approval for each wave of data collection was obtained from the Faculty of Health Sciences Research Ethics Committee at Trinity College Dublin, Ireland. All participants provided written informed consent.

Data collection involved a computer-assisted personal interview (CAPI) with a trained interviewer in the participant’s home, as well as a self-completion questionnaire. As part of the CAPI, TILDA participants were asked if they availed of several named services when visiting their pharmacy in the last 12 months. These included: requesting advice about medication; vaccination; blood pressure (BP) or cholesterol checks; advice on smoking cessation or weight management; and diabetes, asthma, or allergy tests. A full list of pharmacy services included in the CAPI questionnaire is included in Appendix 1; medication dispensing was not asked about as a service.

Factors were identified based on the literature and clinical experience which could plausibly be associated with pharmacy services use. These included socio-demographic factors, age, sex, and level of educational attainment (no or primary education, secondary education, or third-level education). Healthcare use factors included the level of GP utilisation (self-reported number of GP visits in the previous 12 months, divided into groups based on quintile), private health insurance status, and their public health cover entitlement. This was categorized as: those with a medical card (with eligibility based on age and household income, covering a third of the population and two-thirds of those aged ≥65 years(9), entitling individuals to a range of public health services at low or no cost), a GP visit card (with eligibility based on a higher income threshold, entitling to free GP visits), or neither of these. TILDA participants are asked to report doctor-diagnosed health conditions (based on a list of common conditions, and with the opportunity to provide “other” responses) and medications they take on a regular basis. The latter was used to determine participants with polypharmacy (≥5 medications, excluding supplements) and major polypharmacy (≥10 medications). Reported medications were also used to identify participants taking high-risk medication classes (anticoagulants, NSAIDs, opioids, diuretics, antiplatelets, antimicrobials, insulin and hypoglycemics).(10) Last, within the self-completion questionnaire, participants were asked how often they felt lonely. In Ireland, loneliness has been shown to be positively associated with the number of general practitioner visits(11); however, the relationship between loneliness and pharmacy service use in Ireland has not been described.

All study analysis were performed using Stata 17 (StataCorp. 2021. *Stata Statistical Software: Release 17*. College Station, TX: StataCorp LLC). The characteristics of all included participants were described. The prevalence of reported use of any of the above services, as well as the prevalence for each service separately, were summarised among all participants. The relationship between high-risk medication categories and the pharmacy service of requesting advice about medications was assessed using chi-square tests. The relationship between the above characteristics and reporting use of any pharmacy services was assessed using multivariate logistic regression, generating adjusted odds ratios with 95% confidence intervals (CI). A further regression model was fitted with requesting advice about medications as the dependent variable (as the most prevalent service reported). Statistical significance was assumed at p<0.05.

## Results

This study included 5,782 participants, 55.5% were female with a mean age of 68 years. A total of 96.6% of participants (5,587) reported visiting a pharmacy in the previous 12 months, and almost one fifth of these (1,094, 18.9%) availed of at least one specified pharmacy service. A summary of the participants characteristics and a comparison of characteristics between service non-users and users is included in Table 1.

**Table 1.** Descriptive statistics for overall participant demographics and comparison of participants between any pharmacy service non-users and users (n = 5,782).

| Variable | Total<br>n (%)<br>(n = 5782) | No Pharmacy Service Use<br>n (%)<br>(n=4688) | Any Pharmacy Service Use<br>n (%)<br>(n=1094) |
| --- | --- | --- | --- |
| **Age group** |  |  |  |
| 56-64 years | 2,313 (40.0) | 1,893 (40.4) | 420 (38.4) |
| 65-74 years | 2,088 (36.1) | 1,674 (35.7) | 414 (37.8) |
| ≥75 years | 1,381 (23.9) | 1,121 (23.9) | 260 (23.8) |
| **Female sex** | 3,212 (55.6) | 2,531 (54.0) | 681 (62.3) |
| **Education** |  |  |  |
| Primary/none | 1,360 (23.5) | 1,151 (24.6) | 209 (19.1) |
| Secondary | 2,294 (39.7) | 1,902 (40.6) | 392 (35.8) |
| Third/higher | 2,128 (36.8) | 1,635 (34.9) | 493 (45.1) |
| **Health Cover** |  |  |  |
| None | 2,471 (42.8) | 2,033 (43.4) | 438 (40.1) |
| Medical card | 2,681 (46.4) | 2,194 (46.8) | 387 (44.6) |
| GP visit card | 626 (10.8) | 459 (9.8) | 167 (15.3) |
| **Private health insurance** | 3,506 (60.7) | 2,790 (34.5) | 716 (65.5) |
| **Number of regular medications, mean (SD)** | 2.97 (2.87) | 2.82 (2.85) | 3.56 (2.94) |
| **Taking ≥5 medications** | 1,487 (25.7) | 1,121 (23.9) | 366 (33.5) |
| **Taking ≥10 medications** | 190 (3.3) | 143 (3.0) | 47 (4.3) |
| **Frequency of loneliness** |  |  |  |
| Rarely or never | 4,600 (79.7) | 3,785 (80.9) | 815 (74.5) |
| Some of the time | 725 (12.6) | 555 (11.9) | 170 (15.5) |
| Moderate amount | 343 (5.9) | 261 (5.6) | 82 (7.5) |
| All of the Time | 103 (1.8) | 76 (1.62) | 27 (2.47) |
| **Taking a high-risk medication** | 2,444 (42.3) | 1,908 (40.7) | 536 (49.0) |
| **Diabetes** | 532 (9.2) | 417 (8.9) | 115 (10.5) |
| **High cholesterol** | 2,139 (37.0) | 1,677 (35.8) | 462 (42.2) |
| **High blood pressure** | 2,894 (50.0) | 2,287 (48.8) | 607 (55.5) |
| **Other CV condition** | 491 (8.5) | 372 (7.9) | 119 (10.9) |
| **Respiratory condition** | 650 (11.2) | 477 (10.2) | 173 (15.8) |

The most common services reported were requesting advice about medications (786, 13.6%), blood pressure monitoring (184, 3.2%), and vaccination (166, 2.9%). A summary of frequency of service use across all participants is included in Figure 1. Compared to those not using any services, service users were a similar age (mean 68 years), but were taking more medications (mean 3.6 versus 2.8), were more often female (64.1% versus 54.2%), had higher educational attainment, and had higher GP visit rates.

**Figure 1.**
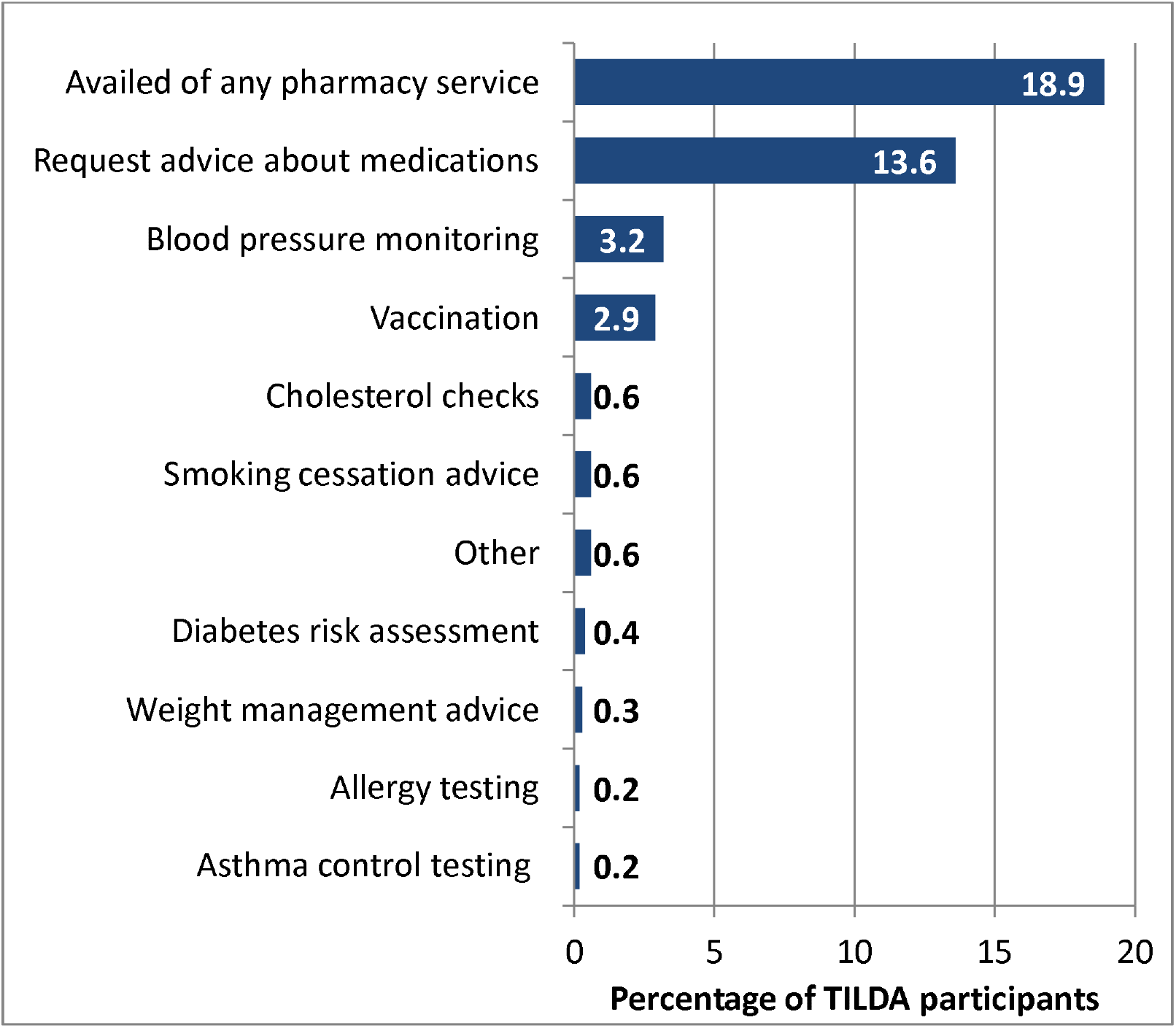
Overall frequency of community pharmacy service use reported by TILDA participants.

Considering the relationship between specific high-risk medications and requesting advice about medications, there was a statistically significant associations for anticoagulant, antiplatelet, and NSAID medication use (Figure 2). Further, the composite for any high risk medication use was associated with requesting advice about medications. Results from Chi-square tests for high risk medication category and requesting advice about medications at the community pharmacy are included in Appendix 2.

**Figure 2.**
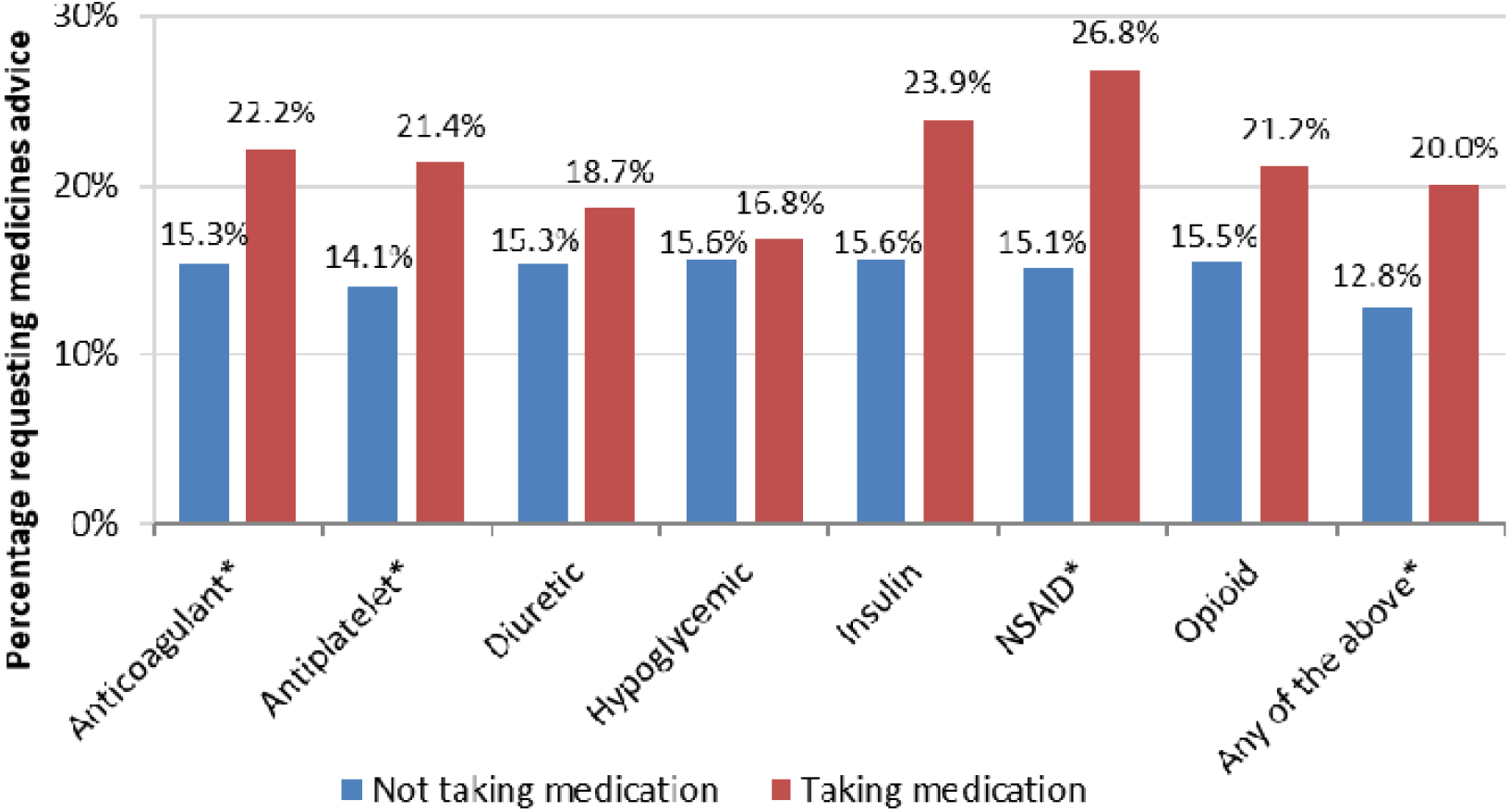
Prevalence of requesting advice about medications by use of high-risk medication classes. * denotes a statistically significant difference.

Controlling for other factors, the following were associated with a higher likelihood of availing of pharmacy services: female sex (adjusted OR 1.32, 95%CI 1.14-1.52), third level education (OR 1.85, 95%CI 1.51-2.27), higher rates of GP visits, private health insurance (OR 1.29, 95%CI 1.07-1.56), higher number of medications, loneliness, and a diagnosed respiratory condition (OR 1.42, 95% CI 1.14-1.74). A complete summary of service use odds ratios and 95% confidence intervals is included in Figure 3. The relationship between these factors and requesting medicines advice were similar (Appendix 3), and notably taking a high-risk medication was significantly associated (OR 1.24, 95%CI 1.01-1.52).

**Figure 3.**
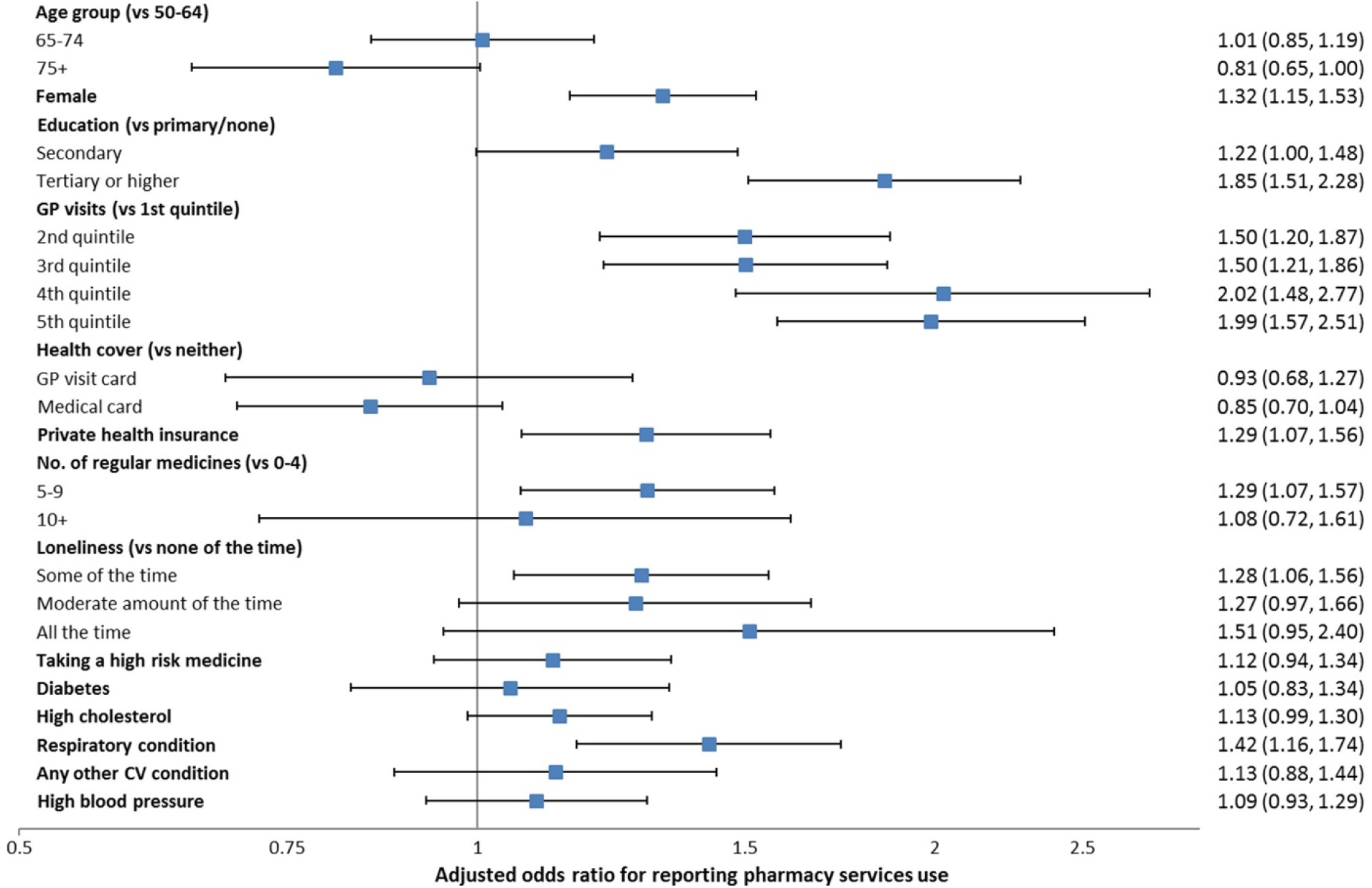
Factors associated with reporting any pharmacy services use in multivariate logistic regression.

## Discussion

This study identified that nearly one-fifth of individuals who visited a pharmacy in the past 12 months used a non-dispensing pharmacy service. The most common service reported was requesting advice about medications. Service use outside of medication advice was relatively low, with the highest services used being blood pressure monitoring (3.2%) and vaccination (2.9%). These findings reflect those in existing community pharmacy literature. First, a number of studies have focused on community pharmacist perspectives on enhanced services, with pharmacists reporting they most frequently provide services considered standard practice while occupying conventional roles within the healthcare system (i.e. medication dispensing).(12-14) There is wide variety in pharmacist willingness to provide and provision of pharmacy services across and between countries(15), with countries like New Zealand effectively scaling select pharmacy services (e.g. provision of trimethoprim for uncomplicated urinary tract infections, 85% of pharmacies surveyed) while other services are less frequently reported (e.g. supply selected oral contraceptives, 44% of pharmacists surveyed).(16) Although pharmacists in other countries report willingness to provide enhanced or expanded pharmacy services, information on the extent of service offering, availability, and uptake is limited.(17, 18)

Further, lack of patient awareness of pharmacy services may contribute to low uptake and repeated use. A systematic review of patient and public perspectives of community pharmacy services in the United Kingdom identified that low public or patient awareness of pharmacy services was a common theme in existing literature, with lack of publicity and exposure to pharmacy services identified as potential explanations.(19) Further, a patient preference study by Janet et. al. included in the systematic review identified that 28 participants (15.8%) chose a pharmacy providing extended services as their least preferred pharmacy, compared to just one patient who selected the pharmacy as their first choice.(19, 20) The review also identified that patient perceptions of the role of pharmacists and physicians were contributing factors to service use, with a number of facilitators and barriers to service use identified.(19) Newer studies have focused on patient preferences for enhanced services provided in the community pharmacy setting using discrete choice experiments, measuring patient preference with utility.(21, 22) A study examining patient preferences for enhanced services in the community pharmacy setting found that services focusing on the provision of prescription drugs for minor ailments, pharmacogenetic testing, and point-of-care testing resulted in negative utility values, suggesting that older patients taking more medications expressed disfavor in the provision of these services and may prefer pharmacies who offer standard pharmacy services.(23) Conversely, a study exploring expanded pharmacy service use in Australia identified that the mean number of pharmacy services that patients felt their community pharmacists should provide was significantly higher for older patients (>55) compared to those who were less than 25 years of age.(24) Overall, the evidence for patient preference and expectation for pharmacy services is mixed, with these differences potentially explained by a difference in the type of pharmacy services offered and included in the studies and patient-specific characteristics like gender and education level.

In addition to service availability, patient awareness, and preferences for pharmacy services, patient-specific factors have been shown to contribute to service use. In this study, patient-specific factors which were positively associated with service use were female sex, third level education, higher rates of GP visits, private health insurance, higher number of medications, loneliness, and a diagnosed respiratory condition. In the existing literature, number of medications are associated with an increase in service use.(25) A study by Picker et. al. found that the prevalence of 30-day hospital readmissions was associated with patients’ number of discharge medications, with a larger number of medications increasing risk of readmission.(26) In addition to number of medications, this study identified that certain categories of high-risk medications were associated with requesting advice about medications specifically. Additionally, a large body of literature has identified multimorbidity, often defined as the coexistence of two or more conditions, as a predictor of increased health service use.(27, 28) Multimorbidity has been associated with potentially inappropriate prescribing and polypharmacy (taking 5 or more medications).(29-32) As such, the results from this study highlight the relationship between multimorbidity, medications, and pharmacy service use, suggesting that patients who may most benefit from pharmacist advice (i.e. patients with multiple conditions, numerous medications, high-risk medications) are availing of community pharmacy services. Finally, higher rates of GP visits were associated with pharmacy service use, suggesting patients may avail of pharmacy services as a complement, rather than a substitute for visiting their GP.

Importantly, loneliness has been identified as a contributing factor to increased health service use amongst older populations in multiple studies.(33-35) Loneliness, recognised as a significant public health issue, has a number of health and economic implications, including increased risk of cardiovascular disease.(35, 36) Existing literature has identified community pharmacies as a potential health promotion resource for older individuals experiencing loneliness, assisting patients in identifying community resources (e.g. care managers and elderly day cares) and engaging in social prescribing.(35, 37, 38) In recent years, the role of community pharmacists in mental health has continued to expand, specifically in the development and provision of mental health first aid.(39,40) While not specifically listed as a pharmacy service in TILDA, community pharmacy training and interventions focused on psychological well-being and mental health may provide older adults in Ireland with essential psychological support and help to decrease physician visits and healthcare costs.(35)

## Limitations and Future Research

This study has a number of limitations. First, TILDA focuses on middle-aged and older adults (all of whom were 56 years and older at wave 4) in Ireland, limiting the generalisability of the findings to other countries and individuals who fall outside of TILDA inclusion criteria. Additionally, while patients reported a variety of pharmacy service use, information on the range of pharmacy services available in Ireland is relatively limited.(41) Future research should focus on exploring the availability of pharmacy services in Ireland, with specific emphasis on pharmacy services and patient perspectives. Additionally, studies focused on the specific strategies to increase public and patient awareness of existing pharmacy services are needed, emphasising existing facilitators and barriers to enhanced service use. Finally, patients may benefit from studies focusing on the development and implementation of additional pharmacy services addressing loneliness and mental health.

The data in this study were collected before the onset of the COVID-19 pandemic in 2020. The pandemic spurred expansion in the role of community pharmacists internationally and in Ireland,(2,42,43) with growth in additional services offered given the reduced access to other healthcare providers.(42,43) This included delivery of COVID-19 vaccination, and the enhanced awareness of such expanded pharmacist roles among the population may have impacted on the use of pharmacy services generally since 2020. Change in pharmacy services use could be examined in future research analysing data from TILDA participants collected since the onset of the COVID-19 pandemic.

## Conclusions

A high proportion of middle-aged and older adults visit community pharmacy and a fifth avail of specified pharmacy services. Despite advances in the services offered in pharmacies, standard pharmacy services such as medicines advice remains at the core of pharmacists’ practice. Patients on multiple medications or with increased GP visits are more likely to use community pharmacy services, suggesting patients may avail of pharmacy services as a complement, rather than a substitute, to visiting their GP. Community pharmacy services were more often used by people who are lonely, identifying an important opportunity for community pharmacies to develop additional interventions focused on health and social support.

## Data Availability

Information on accessing data from TILDA is available at https://tilda.tcd.ie/data/accessing-data/

## Appendices

### Appendix 1. TILDA CAPI Pharmacy Service Questionnaire Item

In the last 12 months when [you/name] visited the pharmacy did [you/he/she] avail of any of the following services?

1. Request advice about medications
2. Blood pressure monitoring
3. Smoking cessation advice
4. Weight management advice
5. Diabetes risk assessment
6. Asthma control testing
7. Allergy testing
8. Cholesterol checks
9. Vaccination
10. Did not visit pharmacy in the last 12 months
95. Other (please specify) [Go to HU081oth]
96. None of these services
98. DK
99. RF

If Other: (please specify)

### Appendix 2. Relationship between high risk medication and patient requesting advice about medication at a community pharmacy, frequencies and chi-square test p-values

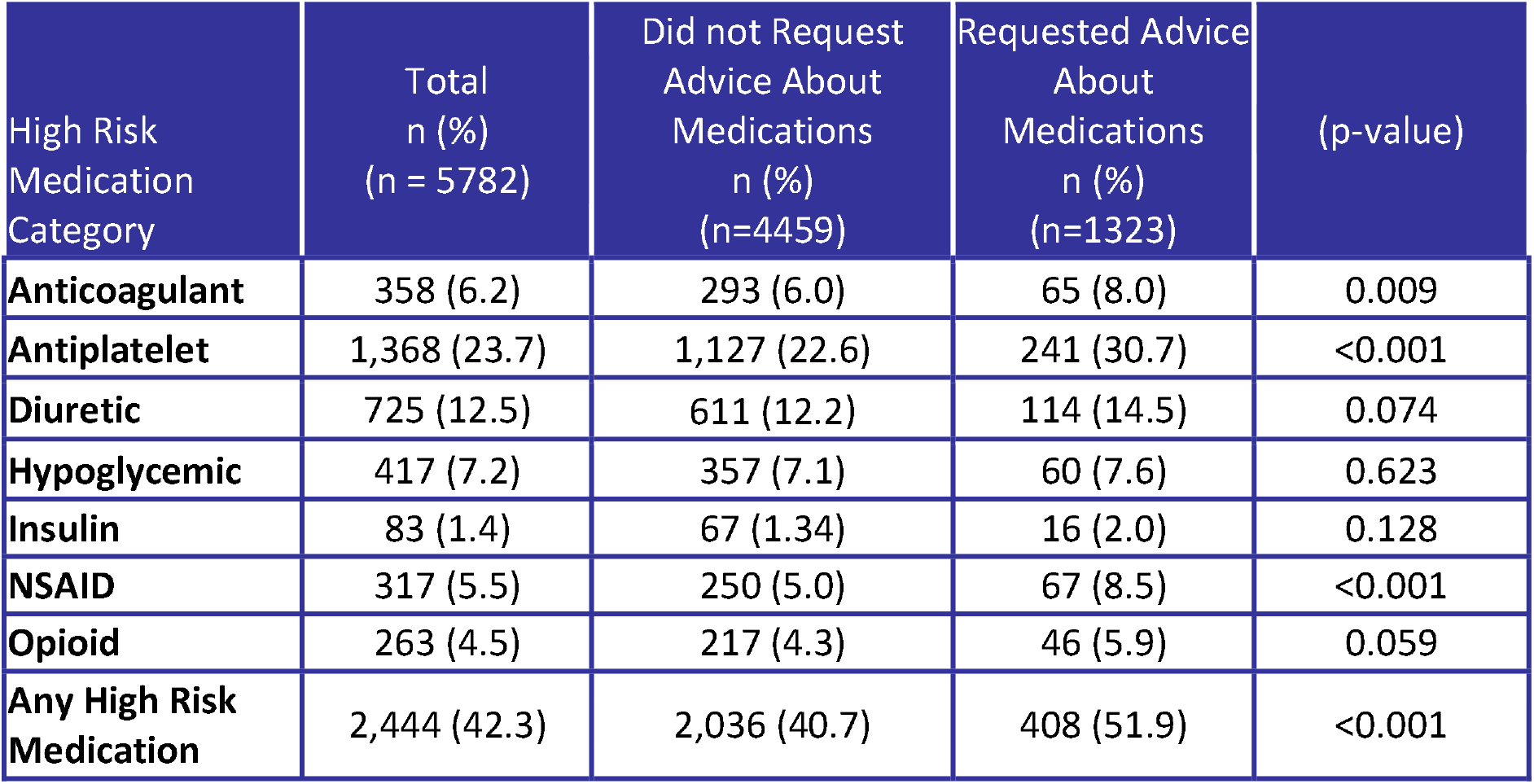

### Appendix 3. Factors associated with reporting requesting advice about medications in multivariate logistic regression

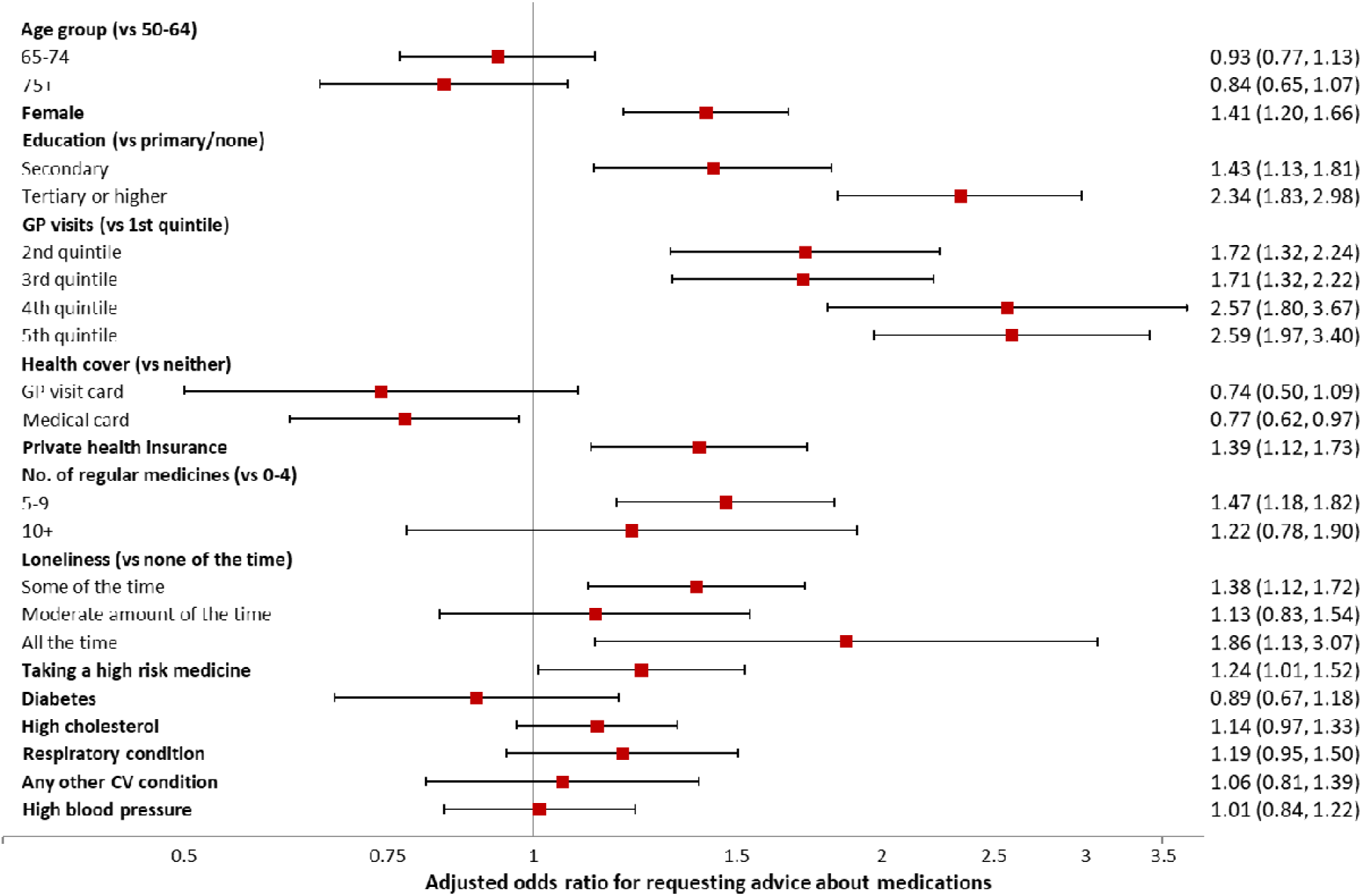

